# Assessment of oxidative stress markers in elderly patients with SARS-CoV-2 infection and potential prognostic implications. An observational study

**DOI:** 10.1101/2022.05.11.22274952

**Authors:** Nestor Vazquez-Agra, Ana-Teresa Marques-Afonso, Anton Cruces-Sande, Ignacio Novo-Veleiro, Antonio Pose-Reino, Estefania Mendez-Alvarez, Ramon Soto-Otero, Alvaro Hermida-Ameijeiras

**Affiliations:** Department of Internal Medicine. University Hospital of Santiago de Compostela, 15706, A Choupana Street, A Coruña, Spain; Laboratory of neurochemistry. Department of Biochemistry and Molecular Biology. Faculty of Medicine. University of Santiago de Compostela, 15782, San Francisco Street, A Coruña, Spain

**Author notes:** Corresponding authors Nestor Vazquez-Agra. Department of Internal Medicine. University Hospital of Santiago de Compostela, 15706, A Choupana Street, A Coruña, Spain. Anton Cruces-Sande. Laboratory of neurochemistry. Department of Biochemistry and Molecular Biology. Faculty of Medicine. University of Santiago de Compostela, 15782, San Francisco Street, A Coruña, Spain. **Author contributions:** Conceptualization – Nestor Vazquez-Agra, Anton Cruces-Sande, Alvaro Hermida-Ameijeiras. Data curation and formal analysis – Ana-Teresa Marques-Afonso, Ignacio Novo-Veleiro, Nestor Vazquez-Agra. Investigation and methodology – Nestor Vazquez-Agra, Anton Cruces-Sande. Supervision and validation: Estefania Mendez-Alvarez, Ramon Soto-Otero, Antonio Pose-Reino, Alvaro Hermida-Ameijeiras. Writing (Original Draft Preparation) – Nestor Vazquez-Agra. Writing (Review & Editing): Nestor Vazquez-Agra. Final approval of the version to be published – All authors.

**Keywords:** Oxidative stress, thiobarbituric acid reactive substances, lipid peroxidation, thiol, SARS-CoV-2, prognostic

## Abstract

The aim of the study was to evaluate the correlation of plasma levels of thiobarbituric acid reactive substances (TBARS) and reduced thiols with morbidity, mortality and immune response in SARS-CoV-2 infection. This was an observational study that included inpatients with SARS-CoV-2 infection greater than 65 years old. Individuals were followed up until 12 months after hospital discharge. Demographic, clinical and laboratory variables were collected. Plasma levels of TBARS and reduced thiols were quantified as a measure of lipid and protein oxidation, respectively. Events of interest (fatal and non-fatal) were quantified at hospital discharge, third, sixth and twelfth-month post-discharge. The outcomes were differences in oxidative stress markers between groups of interest and time to a negative RT-qPCR and to significant anti-SARS-CoV-2 IgM titers. There were 61 patients (57% women) with a mean age of 83 years old. Patients with higher levels of TBARS and lower levels of reduced thiols had more risk of fatal and non-fatal events between admission and the first 12 months post-discharge. The presence of any event (fatal or non-fatal) at the end of the first 12 months post-discharge was correlated with TBARS levels, anti-SARS-CoV-2 IgM titers, lactate dehydrogenase, platelet count and neutrophil and lymphocyte count. We found a correlation between plasma reduced thiols and time to achieve significant anti-SARS-CoV-2 IgM titers. Assessment of some parameters related to oxidative stress could help to identify groups of patients with a higher risk of morbidity and mortality during and after SARS-CoV-2 infection.

## Introduction

The Severe Acute Respiratory Syndrome Coronavirus 2 (SARS-CoV-2) was identified as the cause of a cluster of pneumonia cases in Wuhan (China) at the end of 2019. Symptoms appear in six out of ten patients and about 15% of individuals suffer from severe disease defined as appearance of dyspnea, hypoxemia and major lung involvement on imaging tests [1, 2].

Multiple studies support the existence of a dual interaction between inflammation and oxidative stress witch could leads to an unfavorable inflammatory status that would be related to organ damage and poor prognosis in several acute and chronic diseases [3, 4].

In this line, literature suggests that an exacerbated inflammatory response in predisposed individuals could be one of the main causes of prognostic differences between patients with SARS-CoV-2 infection. Indeed, patients with elevated inflammatory markers such as C-reactive protein (CRP), ferritin, fibrinogen and some pro-inflammatory cytokines, such as interleukin 6 (IL-6) and tumor necrosis factor alpha (TNF-α) are at increased risk of severe complications [5, 6].

However, the role of redox imbalance in the prognosis of SARS-CoV-2 infection in the short and even medium to long term remains poorly understood since most of research on SARS-CoV-2 and oxidative stress are reviews of clinical, translational and base studies related to SARS-CoV-2 virulence and pathogenicity or to several diseases that could share some pathogenic mechanisms [7, 8].

The instability and high reactivity of reactive oxygen species (ROS) implies the need for quantification of secondary but more stable organic products derived from their oxidative action. Some of the most quantified oxidative stress markers in literature are thiobarbituric acid reactive substances (TBARS) and reduced thiols as a measure of lipid and protein oxidation, respectively. The assessment of TBARS and reduced thiols as a prognostic tool is widely supported in some diseases and it would not be negligible that they could also provide prognostic information in SARS-CoV-2 infection [9-11].

Thus, the aim of our study was to quantify plasma TBARS and reduced thiol levels in patients with SARS-CoV-2 infection and to assess whether there is a correlation between these oxidative stress markers and some prognostic variables related to morbidity, mortality and immunity response during and after SARS-CoV-2 infection.

## Material and Methods

### Study design and framework

This was an observational study conducted in a SARS-CoV-2 inpatient unit belonging to the Department of Internal Medicine of the University Hospital of Santiago de Compostela (Galicia, Spain) from November/2020 to January/2021. Patients were recruited at hospital admission and followed up until twelve months after hospital discharge. The events of interest were quantified at hospital discharge, third, sixth and twelfth-month of follow-up.

### Participants

We identified patients older than 65 years old with confirmed SARS-CoV-2 infection by microbiological criteria who met admission criteria (presence of risk factors or severe disease characterized as the appearance of dyspnea, hypoxemia [O2 saturation lower than 94%] or pulmonary involvement greater than 50%). Patients were randomly selected from the complete cohort of patients admitted to hospital with SARS-CoV-2 infection. Individuals with a Barthel index lower than 35 points and those without a record of a laboratory test within the first seven days of admission were excluded [12].

### Clinical variables

All patients were assessed for demographic characteristics (age and sex), cardiovascular (CV) risk factors including smoking status (non-smokers versus current or former smokers), alcohol intake (no consumption versus consumption of any amount), body weight (normal weight versus obesity according to body mass index [BMI]), diabetes mellitus (DM), hyperlipidemia (HLP) and arterial hypertension (AHT). AHT and HLP were defined according to ESC Clinical Practice Guidelines [13, 14]. DM was considered according to American Diabetes Association guidelines (ADA) [15].

Patients were also investigated for the presence of chronic CV disease (coronary arterial and cerebrovascular disease, hearth failure [HF] and atrial fibrillation [AF]), acute or chronic renal impairment and chronic respiratory diseases including chronic pulmonary obstructive disease (COPD), obesity-hypoventilation syndrome and sleep-apnea syndrome. Definitions were taken into account in accordance with the corresponding clinical guidelines [16-18].

The presence of cognitive impairment was confirmed with data collected from the patient’s clinical history and in those cases without previous information we performed the Pfeiffer test [19]. Barthel index punctuation equal or lower than 55 points was considered as moderate physical dependence.

We considered as drug-related variables chronic treatment with anticoagulant, antiplatelet and systemic steroid agents, as well as acute management with antibiotics (azithromycin or others such as B-lactams), systemic steroids and need for supplemental oxygen therapy.

### Laboratory variables

All patients underwent a blood test between days 5 and 7 of hospital admission. Blood samples were obtained at 08:00 AM following overnight fasting. The collected analytical parameters were complete blood count including hemoglobin (Hb), platelet count (PTC) and white blood cell (WBC) count, biochemistry parameters including serum glucose, creatinine, urea and triglycerides (TG), and those related to coagulation. As inflammatory markers they were included the ultrasensitive C-reactive protein (US-CRP), procalcitonin, IL-6, lactate dehydrogenase (LDH), fibrinogen and ferritin [20-22].

Sample collection for the diagnosis of SARS-CoV-2 was performed in the upper respiratory tract in all patients. Detection of SARS-CoV-2 was carried out by using the real-time reverse transcriptase-polymerase chain reaction (RT-qPCR) and results were qualitatively reported as negative, positive or indeterminate. The detection of anti-SARS-CoV-2 IgM antibodies was performed by enzyme-linked immunosorbent assay (ELISA) and results were provided quantitatively [23, 24].

### Assessment of oxidative stress markers

Blood samples were collected in tubes with EDTA, centrifuged in less than 1 hour after extraction at 1000 G and 4°C for 10 minutes with extraction of the plasma fraction and deposited at -80°C for less than 1 month until analysis [25].

TBARS measurement is a well-established method for screening and monitoring lipid peroxidation. There are multiple lipid peroxidation products of which the most relevant and quantified is malondialdehyde (MDA). The MDA-Thiobarbituric acid (TBA) adducts formed by the reaction of MDA and TBA under high temperature (90-100 °C) and acidic conditions were spectrophotometrically measured at 530-540 nm. Assessment of lipid peroxidation was carried out following the protocol of Ohkawa et al [26]. Absorbance was directly proportional to the level of lipid peroxides in plasma and TBARS concentration (expressed in terms of MDA) was given in micromolar (μmol/L). According to literature, physiological concentrations of plasma TBARS range between 0.26 and 3.94 μmol/L [27].

Assessment of reduced thiols is a well-systematized technique for quantification of protein oxidation. Ellman’s technique uses 5,5-dithio-bis-(2-nitrobenzoic acid) as a reagent to form a compound with the sulfhydryl groups of some amino acid residues in proteins giving rise to a colorful pigment that absorbs a wavelength of 412 nm and whose absorbance is proportional to the level of reduced thiols in plasma proteins. Reduced thiols levels were given in millimolar (mmol/L). According to literature, physiological concentrations of plasma reduced thiols range between 0.4 and 0.6 mmol/L [10, 28].

All samples were analyzed in duplicate and a standard calibration for each protocol was performed obtaining a linear model with a coefficient of determination (R2) greater than 98%.

### Outcomes

Mortality was defined as a fatal event. We grouped it into four main categories as follows: I) In-hospital fatal events; II) 3°-month post-discharge fatal events; III) 6°-month post-discharge fatal events and IV) 12°-month post-discharge fatal events.

Non-fatal events were grouped into four main categories as follows: I) In-hospital non-fatal events (which referred to the need for transfer during admission to a critical respiratory care unit); II) 3°-month post-discharge non-fatal events; III) 6°-month post-discharge non-fatal events; IV) 12°-month post-discharge non-fatal events (which referred to the need for readmission to hospital for respiratory or cardiovascular disease within 3, 6 and 12 months post-discharge).

Total events were defined as presence or absence of any fatal or non-fatal events for each of the groups listed previously (In-hospital, 3°, 6° and 12°-month post-discharge). The other outcomes were time to significant anti-SARS-CoV-2 IgM titers (considering mean titers as the threshold) and time to a negative RT-qPCR.

### Calculation of sample size and treatment of variables

For an unknown population size, considering a variance of 0.5 and 0.05 with a threshold for the mean difference to be detected of 0.5 and 0.05 units for TBARS and reduced thiols respectively, the sample size calculated to estimate the mean difference between groups with a 95% confidence interval was a minimum of n = 30 patients [29]. Variables were collected according to data provided by the regional digital health records (IANUS) belonging to the Galician (Spain) Health Service (SERGAS). Most of the clinical variables were coded as qualitative ones and laboratory parameters including TBARS and reduce thiols were collected as continuous quantitative variables. Outcome variables were coded as dichotomous qualitative variables.

### Ethical approval

The study was approved by the Research Ethics Committee of Santiago de Compostela-Lugo (2020/578). All procedures were in accordance with the ethical standards of the responsible committee on human experimentation and with the Helsinki Declaration of 1975. Informed consent was obtained from all patients for being included in the study.

### Statistical analysis

Statistical analysis was performed by using SPSS 22.0 statistical software (SPSS Inc, Chicago, IL). First, we made a descriptive analysis in which frequencies of qualitative variables were expressed as number (n) and percentage (%). The Kolmogorov-Smirnov test was used to determine whether quantitative continuous variables were normally distributed. Normally distributed variables were expressed as mean (m) and standard deviation (± SD) and non-normally ones were expressed as median and interquartile range (IQR). A missing value analysis was carried out on those variables with more than 5% of missing values.

We carried out a comparative analysis between groups of patients according to the outcome variables. The chi-square test was used to compare categorical variables while quantitative variables were compared by using the Student’s t-test or Mann–Whitney U test where appropriate. For multivariate analysis, a binary logistic regression model was developed on the relevant clinical and laboratory variables. The calculated probability was evaluated with a receiver operating characteristic (ROC) curve model. Time to significant anti-SARS-CoV-2 IgM titers and time to a negative RT-qPCR were used to construct a survival analysis model by using the Kaplan–Meier estimator attending to the level of TBARS and reduced thiols. A P-value less than 0.05 was considered for statistical significance.

## Results

Quantitative variables were normally distributed and we only registered more than 5% of missing values in alcohol intake, smoking habit (current/former smokers) and level of physical dependence (Barthel index), which were found to be missing values at random. There were 61 patients (57% women) with a mean age of 83 years old who were included in the study, and among them, 42 (69%), 31(51%) and 15(25%) suffered from AHT, HLP and DM respectively. A total of 27(44%) and 21(34%) individuals were affected of HF and AF. Approximately one out of four individuals had tobacco or alcohol abuse and most of them were former users (data not shown). The mean levels of TBARS and reduced thiols were 3.11 ± 0.09 μmol/L and 0.46 ± 0.07 mmol/L respectively. The mean levels of anti-SARS-CoV-2 IgM titers were 40 ± 38 U/mL. General results are shown in Table 1 and complete clinical features are provided in S1 Table.

**Table 1.**
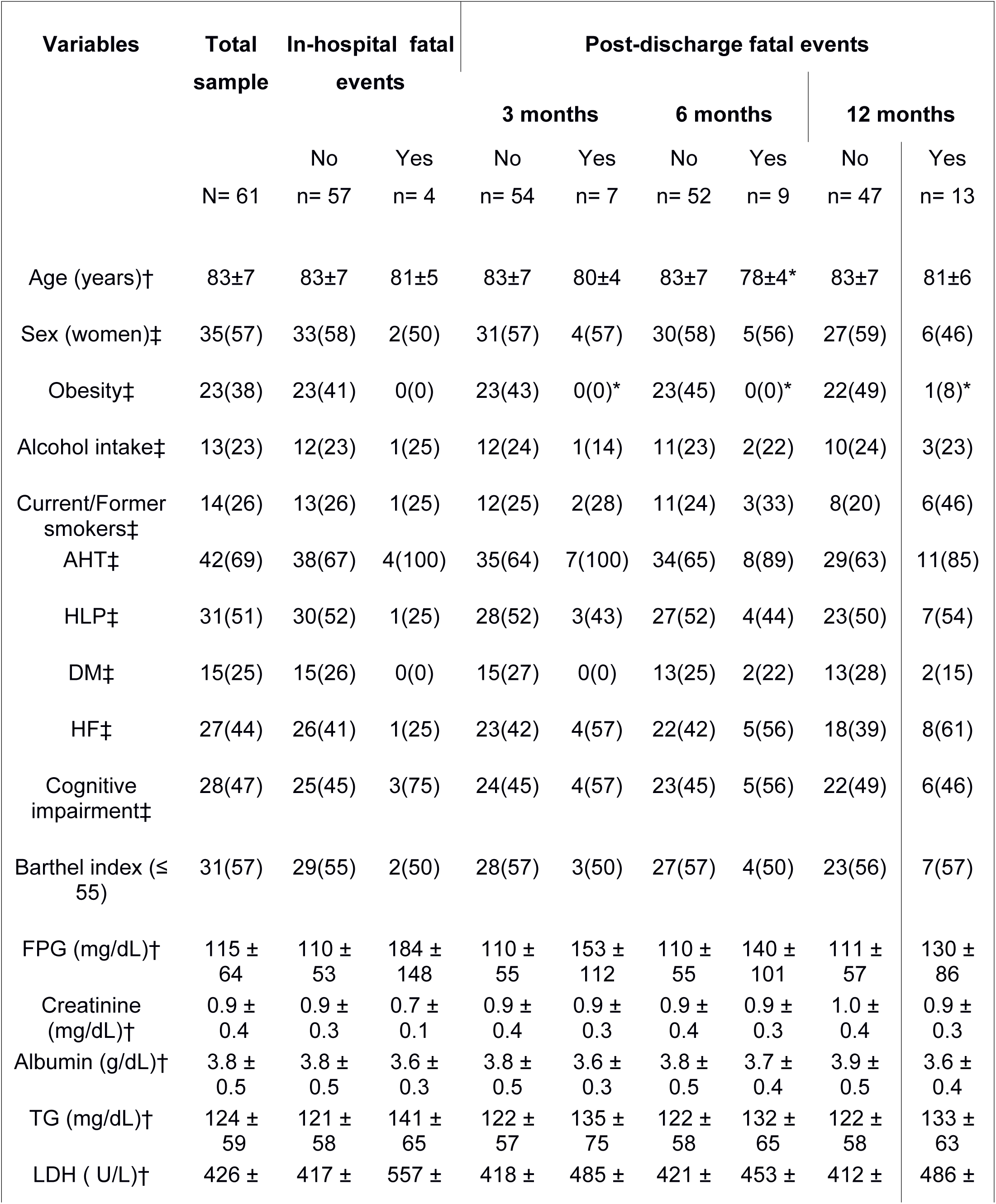
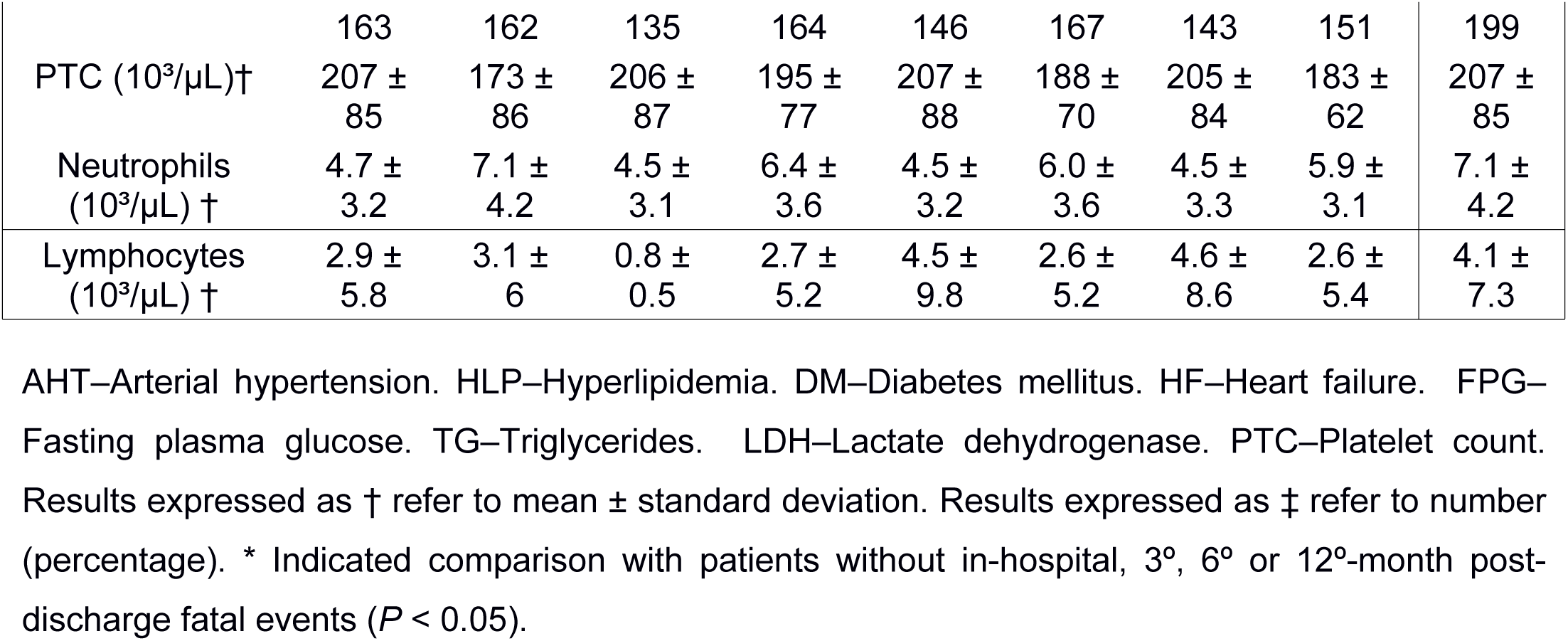
Clinical and laboratory features of comparison groups attending to mortality.

### Fatal events

Results of the comparison groups are shown in Table 1 and extended in S1-2 Tables. In general terms, younger patients, with lower body weight and greater use of antibiotics or systemic steroids during admission presented a greater number of fatal events with statistically significant differences in the groups of 3°, 6° and 12°-month post-discharge fatal events. Regarding laboratory variables, there were no relevant differences between the comparison groups.

Patients who presented a fatal event had higher TBARS levels than controls measured in μmol/L and expressed as m ± SD. These differences reached statistical significance in the following groups: In-hospital (no: 3.02 ± 1.01, yes: 4.40 ± 1.52; P= 0.014) and 6°-month post-discharge fatal events (no: 2.99 ± 0.98, yes: 3.81 ± 1.44; P= 0.038). The levels of reduced thiols measured in mmol/L and expressed as m ± SD were lower in patients who suffered a fatal event. Such differences reached statistical significance in the following groups: 6°-month (no: 0.47 ± 0.07, yes: 0.41 ± 0.05; P= 0.040) and 12°-month (no: 0.47 ± 0.07, yes: 0.42 ± 0.06; P= 0.032) post-discharge fatal events. The results are extended in Fig 1A and 1B.

**Fig 1.**
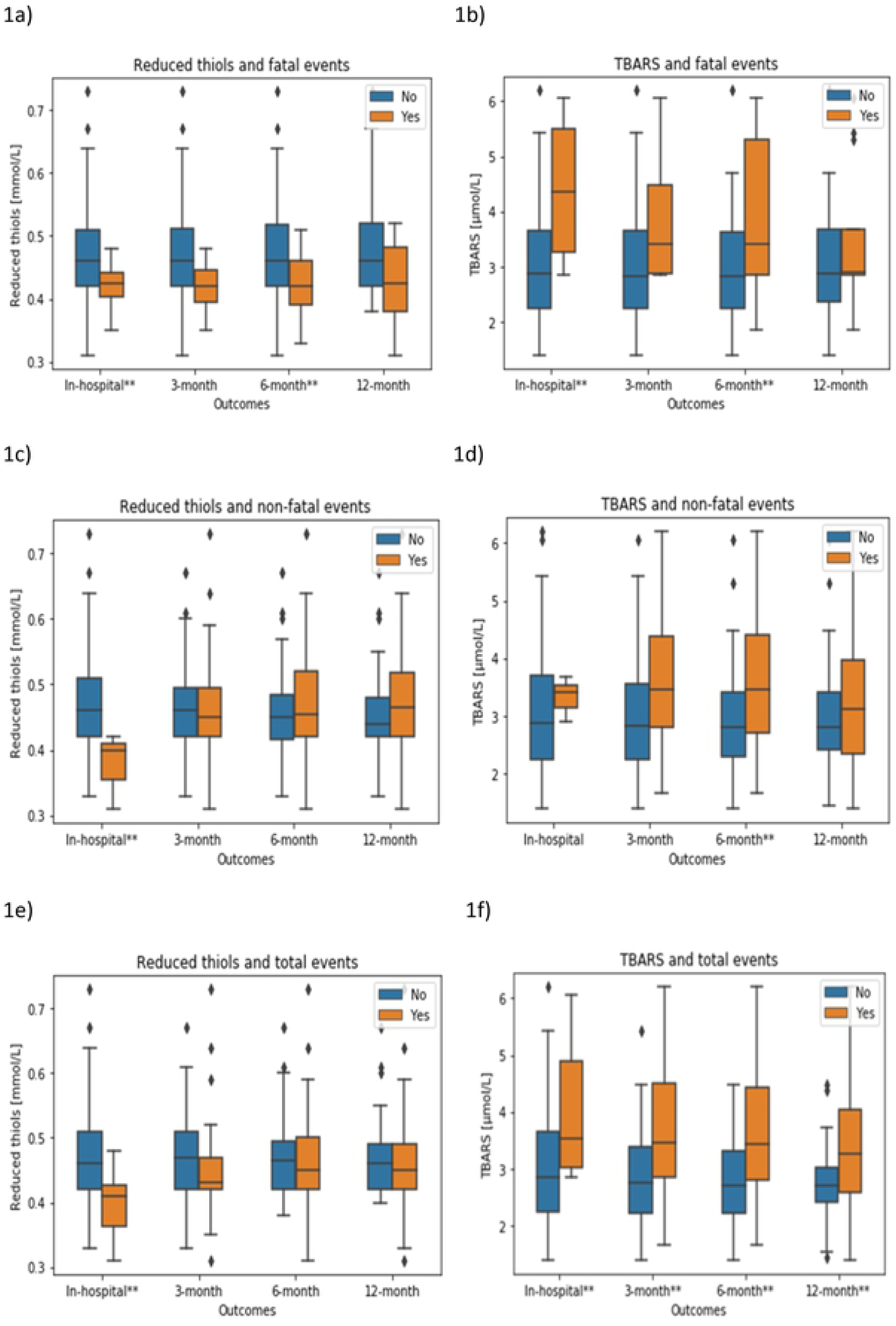
Difference between plasma levels of TBARS and reduced thiols in groups of interest. (A-B) In-hospital and post-discharge fatal events. (C-D) In-hospital and post-discharge non-fatal events. (E-F) In-hospital and post-discharge total events. Results are shown in mean ± standard deviation. TBARS–Tiobarbituric acid reactive substances. **Refers to P-value <0.05.

### Non-fatal events

The results of the comparison groups are shown in Table 2 and extended in S3-4 Tables. In general terms current or former smokers presented a higher number of non-fatal events with statistically significant differences in the 6°-month and 12°-month post-discharge non-fatal events groups. Regarding laboratory variables, we only observed relevant differences in LDH and PTC for 6°-month and 12°-month post-discharge non-fatal events respectively.

**Table 2.** Clinical and laboratory features of comparison groups attending to non-fatal events.

| Variables | In-hospital non-fatal events |  | Post-discharge non-fatal events |  |  |  |  |  |
| --- | --- | --- | --- | --- | --- | --- | --- | --- |
|  |  |  | 3 months |  | 6 months |  | 12 months |  |
|  | No<br>n= 58 | Yes<br>n= 3 | No<br>n= 44 | Yes<br>n= 17 | No<br>n= 38 | Yes<br>n= 23 | No<br>n= 28 | Yes<br>n= 33 |
| Age (years)† | 82 ± 7 | 84 ± 6 | 83 ± 8 | 83 ± 5 | 83 ± 8 | 82 ± 6 | 82 ± 7 | 83 ± 8 |
| Gender (women)‡ | 33(57) | 2(67) | 26(59) | 9(53) | 23(61) | 12(52) | 18(64) | 17(52) |
| Obesity‡ | 23(40) | 0(0) | 17(40) | 6(35) | 15(41) | 8(35) | 10(36) | 13(41) |
| Alcohol intake‡ | 13(25) | 0(0) | 8(21) | 5(29) | 6(18) | 7(30) | 4(16) | 9(29) |
| Current/Formersmokers‡ | 13(25) | 1(33) | 8(21) | 6(35) | 5(16) | 9(39)* | 3(12) | 11(37)* |
| AHT‡ | 39(67) | 3(100) | 27(61) | 15(88)* | 24(63) | 18(78) | 18(64) | 24(73) |
| HLP‡ | 29(50) | 2(67) | 21(48) | 10(59) | 18(47) | 13(57) | 13(46) | 18(55) |
| DM‡ | 15(26) | 0(0) | 10(23) | 5(29) | 8(21) | 7(30) | 6(21) | 9(27) |
| HF‡ | 25(43) | 2(67) | 17(39) | 10(59) | 14(37) | 13(57) | 9(32) | 18(55) |
| Cognitive impairment‡ | 27(47) | 1(33) | 21(49) | 7(41) | 19(51) | 9(39) | 14(52) | 14(42) |
| Barthel index (≤ 55) ‡ | 30(57) | 1(33) | 21(55) | 10(59) | 18(56) | 13(56) | 17(76) | 14(44)* |
| FPG(mg/dL)† | 115 ± 65 | 111 ± 14 | 112 ± 67 | 122 ± 56 | 112 ± 71 | 119 ± 50 | 111 ± 62 | 117 ± 66 |
| Creatinine (mg/dL)† | 0.9 ± 0.4 | 0.8 ± 0.2 | 0.9 ± 0.3 | 1.0 ± 0.5 | 0.9 ± 0.3 | 0.9 ± 0.4 | 0.9 ± 0.3 | 1.0 ± 0.4 |
| Albumin (g/dL)† | 3.8 ± 0.5 | 3.4 ± 0.5 | 3.8 ± 0.5 | 3.7 ± 0.3 | 3.8 ± 0.5 | 3.7 ± 0.3 | 3.8 ± 0.6 | 3.8 ± 0.4 |
| TG (mg/dL)† | 120 ± 58 | 182 ± 41 | 123 ± 58 | 124 ± 62 | 124 ± 59 | 122 ± 59 | 124 ± 43 | 123 ± 70 |
| LDH ( U/L)† | 421 ± 165 | 511 ± 91 | 407 ± 150 | 474 ± 187 | 391 ± 127 | 483 ± 198* | 401 ± 125 | 448 ± 189 |
| PTC (10<sup>3</sup>/μL)† | 204 ± 86 | 212 ± 75 | 192 ± 68 | 237 ± 115 | 191 ± 71 | 227 ± 102 | 180 ± 68 | 225 ± 93 * |
| Neutrophils (10<sup>3</sup>/μL) † | 4.7 ± 3.2 | 6.1 ± 2.4 | 4.4 ± 3.1 | 5.5 ± 3.7 | 4.5 ± 3.2 | 5.2 ± 3.3 | 4.6 ± 3.4 | 4.8 ± 3.1 |
| Lymphocytes (10<sup>3</sup>/μL) † | 2.9 ± 5.9 | 3.3 ± 4.6 | 2.3 ± 4.0 | 4.5 ± 8.9 | 2.5 ± 4.3 | 3.7 ± 7.7 | 2.6 ± 4.9 | 3.2 ± 6.5 |

AHT–Arterial hypertension. HLP–Hyperlipidemia. DM–Diabetes mellitus. HF–Heart failure. FPG–Fasting plasma glucose. TG–Triglycerides. LDH–Lactate dehydrogenase. PTC–Platelet count. Results expressed as † refer to mean ± standard deviation. Results expressed as ‡ refer to number(percentage). \* Indicated comparison with patients without in-hospital, 3<sup>o</sup>, 6<sup>o</sup> or 12<sup>o</sup>-month post-discharge non-fatal events ( $P < 0.05$ ).

Patients who suffered from a non-fatal event had higher TBARS levels than controls measured in μmol/L and expressed as m ± SD. Such differences reached relevance at the following groups: 6°-month post-discharge non-fatal events (no: 2.90 ± 1.01, yes: 3.46 ± 1.15; P= 0.052). The levels of reduced thiol groups measured in mmol/L and expressed as m ± SD were lower in patients who suffered non-fatal events. Such differences reached statistical significance in the following groups: In-hospital non-fatal event (no: 0.47 ± 0.07, yes: 0.37 ± 0.05; P= 0.035). The results are extended in Fig 1C and 1D.

### Total events

Patients with any total event had higher TBARS levels than controls measured in μmol/L and expressed as mean (m) ± SD. These differences reached statistical significance in the following groups: In-hospital (no: 3.01 ± 1.02, yes: 4.02 ± 1.33; P= 0.030), 3°-month (no: 2.84 ± 0.91, yes: 3. 63 ± 1.23; P= 0.006), 6°-month (no: 2.7 ± 0.80, yes: 3.51 ± 1.25; P= 0.007) and 12°-month (no: 2.75 ± 0.78, yes: 3.33 ± 1.19; P= 0.043) total events. The levels of reduced thiol groups measured in mmol/L and expressed as m ± SD were lower in patients who suffered a non-fatal event. Such differences reached statistical significance in the following groups: In-hospital total events (no: 0.47 ± 0.07, yes: 0.37 ± 0.05; P= 0.035). The results are extended in Fig 1E and 1F.

Multivariate logistic regression showed a relevant correlation of 12-month total events with TBARS levels, anti-SARS-CoV-2 IgM titers, LDH, PTC and neutrophil and lymphocyte counts. The results of multivariate analysis are shown in Table 3. According to the probability function, a threshold of 0.427 represented a sensitivity and specificity of 80% and 92% for 12°-month post-discharge total events. The results related to the ROC curve over the model are shown in Fig 2.

**Table 3.**
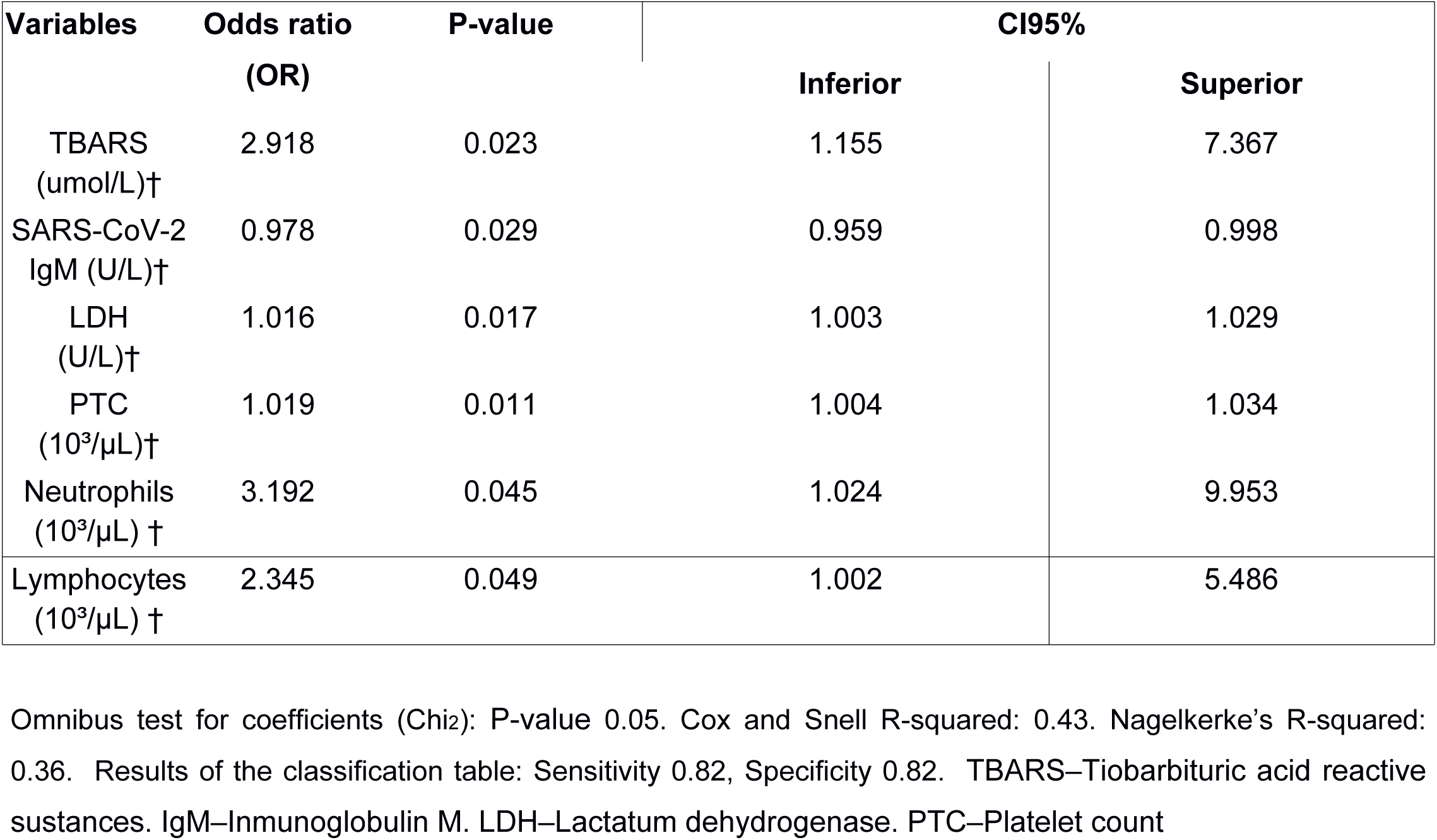
Relevant variables in the model of multivariate logistic regression for the 12°-month post-discharge total events.

| Variables | Odds ratio<br>(OR) | P-value | CI95% |  |
| --- | --- | --- | --- | --- |
|  |  |  | Inferior | Superior |
| TBARS<br>(umol/L)† | 2.918 | 0.023 | 1.155 | 7.367 |
| SARS-CoV-2<br>IgM (U/L)† | 0.978 | 0.029 | 0.959 | 0.998 |
| LDH<br>(U/L)† | 1.016 | 0.017 | 1.003 | 1.029 |
| PTC<br>(10<sup>3</sup>/μL)† | 1.019 | 0.011 | 1.004 | 1.034 |
| Neutrophils<br>(10<sup>3</sup>/μL) † | 3.192 | 0.045 | 1.024 | 9.953 |
| Lymphocytes<br>(10<sup>3</sup>/μL) † | 2.345 | 0.049 | 1.002 | 5.486 |

Omnibus test for coefficients (Chi<sup>2</sup>): P-value 0.05. Cox and Snell R-squared: 0.43. Nagelkerke's R-squared: 0.36. Results of the classification table: Sensitivity 0.82, Specificity 0.82. TBARS–Tiobarbituric acid reactive substances. IgM–Inmunoglobulin M. LDH–Lactatum dehydrogenase. PTC–Platelet count

**Fig 2.**
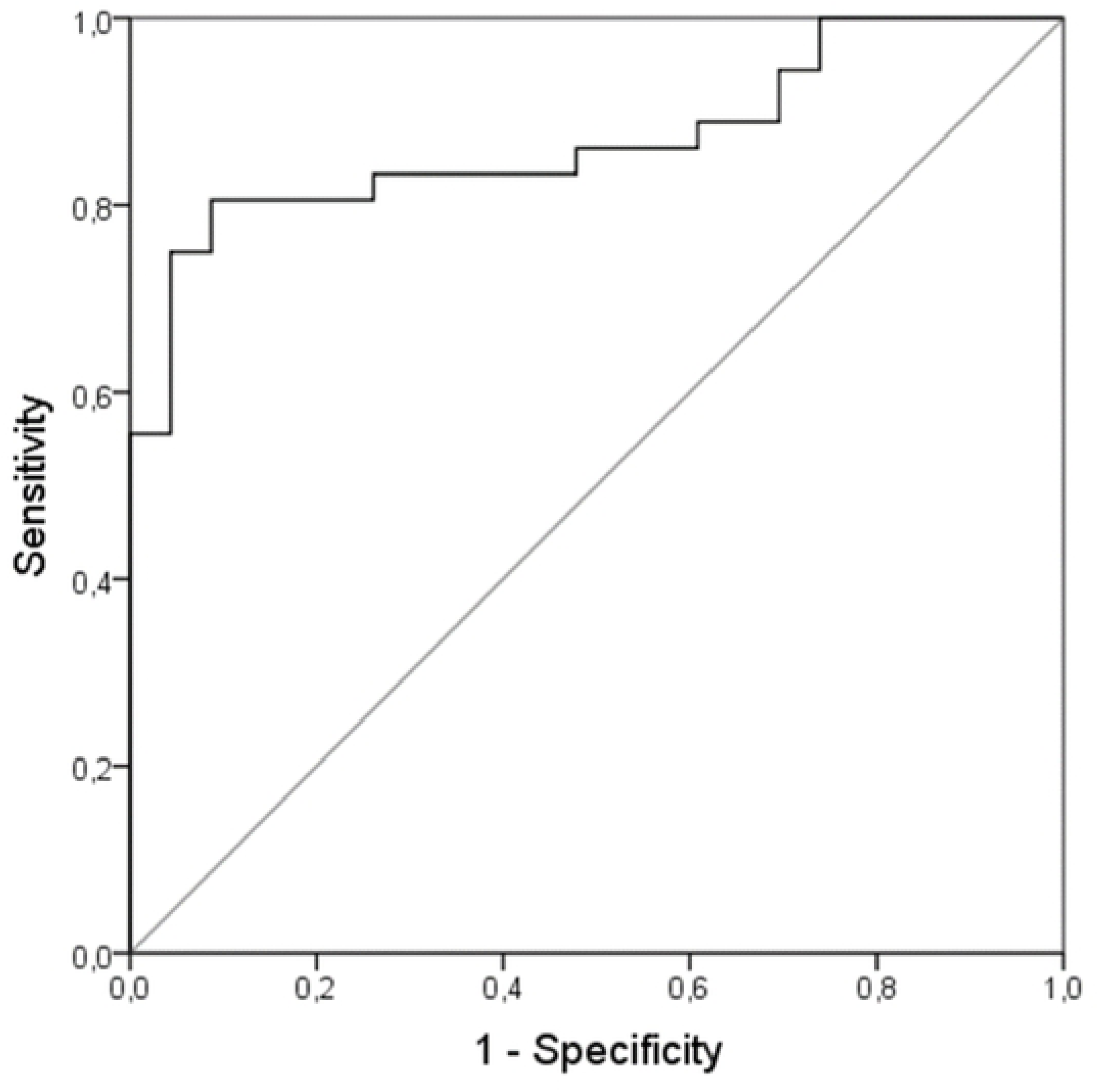
Receiver operating characteristics curve model of some laboratory parameters for the 12°-month post-discharge total events. The variables in the model were plasma TBARS, anti-SARS-CoV-2 IgM titers, LDH, PTC, neutrophils count and lymphocytes count. The results are summarized as follows: Area under curve (AUC) 0.870, standard error (SE) 0.047, P-value <0.001, CI95% 0.778 – 0.961. TBARS–Tiobarbituric acid reactive substances. IgM–Inmunoglobulin M. LDH–Lactate dehydrogenase. PTC–Platelet count.

### Time to a negative RT-qPCR for SARS-CoV-2

No cutoff point for TBARS or reduced thiols levels showed statistical significance for time to a negative RT-qPCR. Figure 3a-b shows the survival analysis using the mean levels of TBARS and reduced thiols as the cut-off point.

**Fig 3.**
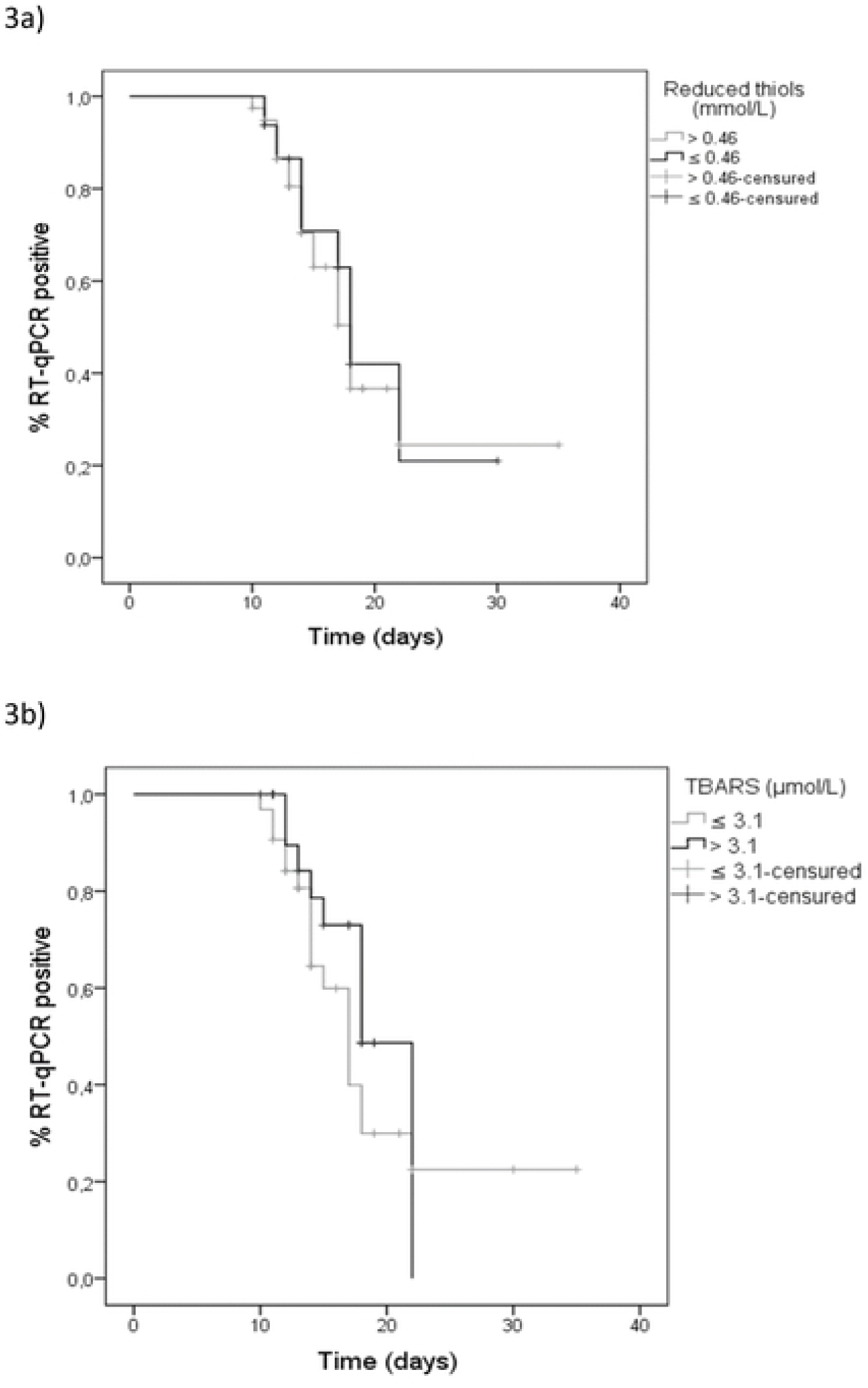
Survival analysis by using the Kaplan–Meier estimator. (A) orrelation of plasma levels of reduced thiols with time to a negative RT-qPCR. Log Rank (Mantel-Cox), p-value= 0.544; (B) Correlation of plasma levels of TBARS with time to a negative RT-qPCR. Log Rank (Mantel-Cox), P-value= 0.675. TBARS–Tiobarbituric acid reactive substances. RT-qPCR– Real-time reverse transcriptase-polymerase chain reaction.

## Time to significant serological titers

Considering a cutoff for significant anti-SARS-CoV-2 IgM titers above the mean (40 U/mL), patients with levels of reduced thiols below and above the mean (0.46 mmol/L) showed relevant differences in time to reach significant anti-SARS-CoV-2 IgM titers. No cut-off point for TBARS levels showed relevant results. Figure 4a-b shows the survival analysis using the mean levels of TBARS and reduced thiols as the cut-off point.

**Fig 4.**
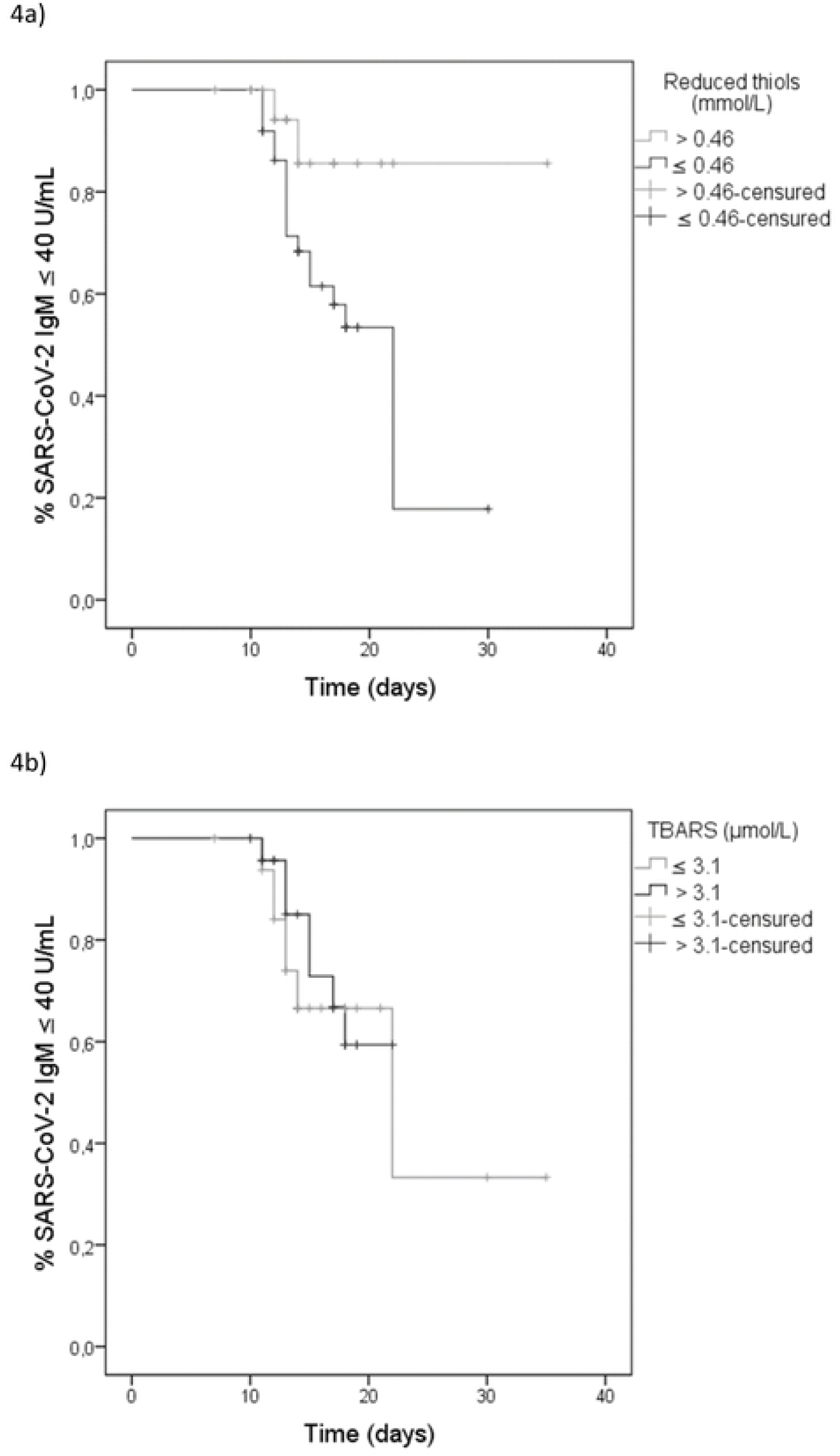
Survival analysis by using the Kaplan–Meier estimator. (A) Correlation of plasma levels of reduced thiols with time to significant anti-SARS-CoV-2 IgM titers. Log Rank (Mantel-Cox), P-value= 0.033. (B) Correlation of plasma levels of TBARS with time to significant anti-SARS-CoV-2 IgM titers. Log Rank (Mantel-Cox), P-value= 0.227. TBARS–Tiobarbituric acid reactive ances.

## Discussion

The most relevant results are summarized as follows: I) Overall, patients with higher levels of TBARS and lower levels of reduced thiols had more risk of fatal and non-fatal events between hospital admission and the first 12 months post-discharge; II) The presence of any event (fatal or non-fatal) at the end of the first 12 months post-discharge was correlated with some analytical parameters including TBARS levels, anti-SARS-CoV-2 IgM titers, LDH, PTC and WBC counts; III) We have not been able to demonstrate a correlation between differences in TBARS and reduced thiols levels and time to a negative RT-qPCR; IV) However, we found a correlation between differences in plasma reduced thiol and time to achieve significant anti-SARS-CoV-2 IgM titers.

In a preclinical setting, many lines of evidence support that a combination between overproduction of ROS and a deprived antioxidant system plays a major role in the pathogenesis of SARS-CoV-2 infection. Experimental animal models of severe acute respiratory syndrome (SARS) attempt to elucidate metabolic pathways that may be involved in redox imbalance during SARS-CoV-2 infection while other studies claim the possible role that certain antioxidant molecules use could play during infection [30, 31].

Metabolic pathways related to oxidative stress could be involved in the severity of SARS-CoV-2 infection in multiple direct and indirect ways. Several studies pointed to the existence of a crosstalk between oxidative stress and inflammation that would involve an interaction between ROS (mainly mitochondrial superoxide and hydroxyl radicals) and some inflammatory mediators due to redox up regulation of some transcription factors such as Nrf2, NF-KB and the NLRP3 inflammasome, as well as their downstream targets, inducing a pro-inflammatory state [32, 33].

However, non-inflammatory cellular pathways related to redox imbalance and their role in SARS-CoV-2 infection severity and prognostic are not yet fully understood. Indeed, if the role that redox status may play in the prognosis of SARS-CoV-2 infection is poorly understood, the consequences of redox imbalance in the medium and long term are still incalculable although it is possible that mortality and admissions one year ahead could be influenced by an unfavorable redox status during the infection.

TBARS are a group of molecules related to lipid peroxidation that were implicated in a multitude of acute and chronic diseases [34]. Literature widely supports the role of lipid peroxidation products in the prognosis of some inflammatory diseases and there are some investigation lines trying to discover metabolic pathways related to lipid peroxidation and its role in the severity and evolution of patients with SARS-CoV-2 infection [35, 36]. Reduced thiols are indicators of protein oxidation and have been widely related to pathogenicity and virulence of SARS-CoV-2 infection and, although there are some studies pointing to a prognostic role of reduced thiols in SARS-CoV-2 infection, scientific evidence is still weak [37, 38].

According to literature, the results related to 12°-month post-discharge total events could suggest at least three concomitant and interrelated processes in SARS-CoV-2 infection with a prognostic role in the medium and long term as follows: I) Impaired oxidative metabolism, tissue hypoxemia and anaerobic metabolism represented by between-group differences in some oxidative stress markers (TBARS) and LDH; II) endothelial dysfunction, platelet activation and a procoagulant state represented by between-group differences in PTC; and III) inflammation and immune activation represented by between-groups differences in neutrophyl and lymphocyte counts, as well as anti-SARS-CoV-2 IgM titers [39-41].

In the end, multiple studies have shown the existence of mechanisms related to redox imbalance and a proinflammatory status in SARS-CoV-2 infection. Some of these studies have also investigated the existence of prognostic differences based on certain inflammatory and oxidative stress markers. Several papers have also focused on the implications of RT-qPCR results and serological titers during SARS-CoV2 infection in terms of mechanisms, duration and prognosis of infection. However, to date there is an evidence gap on the implications that redox status during SARS-CoV-2 infection could have in the medium to long term and we do not know how redox status might affect the time required for a negative RT-qPCR or acquisition of significant anti-SARS-CoV-2 IgM titers. Our study aims to be a humble approximation although further evidence is needed.

### Limitations and strengths

This was a prospective single-center study of real clinical practice carried out in elderly Caucasian patients with SARS-CoV-2 infection from the northwest region of Spain (Galicia) who exhibited similar features than elderly people from the western European countries. Our results should be interpreted with caution when applying them to other population, race or ethnicity.

The sample was not the total population of patients admitted for SARS-CoV-2 in the unit, so the results could be susceptible to a certain sampling error. Although patients were evaluated prospectively, baseline clinical and laboratory data were collected at one point during admission, so some changes in these variables may be relevant and provide more information. Assessing comparability between groups on some variables was complicated because of a low frequency of events of interest in some categories, which could make some results less accurate.

Regarding the results, we found that younger patients with lower body weight had higher mortality, although it is known that they were patients with a higher degree of cognitive impairment who were probably in a poorer nutritional status. On the other hand, current or former smokers presented a higher number of non-fatal events in some comparison groups. However, in most cases the smoking abuse was stopped more than 10-15 years ago, so a strong implication of that differences on the results would be unlikely.

In another vein, the frequency of acute treatments (antibiotics, steroids and oxygen therapy use) was higher in patients with worst outcomes. However, these patients usually had a higher frequency of severe disease, so differences in acute management could be more a consequence of severity rather than adverse prognosis.

## Data Availability

Data cannot be shared publicly in accordance with Article 18.4 of the Spanish Constitution and the Organic Law on Data Protection and Guarantee of Digital Rights (LOPDGDD) of December 6, 2018 the privacy, privacy and integrity of the individual will be protected at all times. Data are available from the regional digital health records (IANUS) belonging to the Galician (Spain) Health Service (SERGAS)/ Research Ethics Committee of Santiago de Compostela-Lugo (2020/578), (contact via telphone number 0034676107918) for researchers who meet the criteria for access to confidential data.

## Acknowledgements

We would like to express our gratitude to the patients who participated in the study and their families for their support. We would like to thank the University Hospital and the University of Santiago de Compostela for the support provided.

## Supporting information

**S1 Table. General features of comparison groups attending to mortality**

**S2 Table. Laboratory findings of comparison groups attending to mortality**

**S3 Table. General features of comparison groups attending to non-fatal events**

**S4 Table. Laboratory findings of comparison groups attending to non-fatal events**

